# Monitoring COVID-19 Occurrence in a Resource-limited Setting – COVID-19 Sentinel Surveillance in Malawi

**DOI:** 10.1101/2024.12.23.24319583

**Authors:** Godwin Ulaya, Alinune Kabaghe, Christel Saussier, Ellen MacLachlan, Joshua Smith-Sreen, Chaplain Katumbi, George Bello, Terence Tafatatha, Limbikani Chaponda, Bernard Mvula, Matthews Kagoli, Benson Chilima, Joseph Bitilinyu-Bangoh, Laphiod Chisuwo, Yusuf Babaye, Moses Chitenje, Barbara Bighignoli, Fred Bangara, Ireen Namakhoma, Annie Chauma-Mwale, Gabrielle O’Malley, Kelsey Mirkovic, Nellie Wadonda-Kabondo

**Author notes:** **Address for correspondence:** Godwin Ulaya, I-TECH Malawi Plot 13/14 1^st^ floor ARWA House, City Centre, Lilongwe. P O Box 30369, Lilongwe Malawi.

## Abstract

The routine COVID-19 surveillance in Malawi that relied on retrospective reporting could not efficiently steer timely measures to the rapidly evolving pandemic. To monitor real-time changes in infections and inform the COVID-19 response, we implemented an active sentinel surveillance system from July to December 2022. SARS-CoV-2 symptomatic and asymptomatic patients in selected health facilities (HFs) and anyone aged ≥5 years entering at Point of Entry (PoEs) sites were eligible to participate. Self-reported epidemiological and clinical data, and nasopharyngeal specimens were collected from 9,305 participants. A higher overall SARS-CoV-2 RT-PCR positivity rate was observed at HFs, 8.9% among symptomatic and 6.5% among asymptomatic patients, versus 3.5% at PoEs. The positivity trends among symptomatic and asymptomatic patient groups showed a similar pattern throughout the period. This active surveillance complemented routine surveillance, especially during a low incidence period and highlighted the need to target both symptomatic and asymptomatic population.

## Introduction

SARS-CoV-2 infection was first confirmed in Malawi on 2nd April 2020 (1). The Government of Malawi subsequently activated a national Public Health Emergency Operations Center to coordinate the multisectoral response to the COVID-19 pandemic (2). Despite the government’s initial response within the capacity of its resources, the surge of cases in the communities and the porous Points of Entry (PoEs) posed challenges to track changes in the incidence of COVID-19. Additionally, the limited investment in and prioritization of public health programs contributed to inadequate capacity in some health facilities (HFs) to routinely test for SARS-CoV-2. As a result, there was a reduced ability to detect cases and monitor changes in real-time (3). Although suspected COVID-19 cases were confirmed using both rapid diagnostic and polymerase chain reaction (PCR) testing and reported, surveillance was passive and circulating variants were not systematically tracked. The testing algorithm used in Malawi was restricted to those that were symptomatic, despite evidence of transmissibility of laboratory-confirmed SARS-CoV-2 infections among asymptomatic individuals (4).

Although preventive and mitigation measures were recommended, periods of low reported COVID-19 incidence were accompanied with a noticeable general decline in adherence to these measures within communities and among healthcare workers. There was also relaxation in the overall routine surveillance system, with reduced testing and delayed reporting. Moreover, the emergence of the polio and cholera outbreaks in early 2022 compounded the situation and diverted attention and resources away from passive COVID-19 surveillance, further hindering the monitoring of the COVID-19 situation in Malawi (5,6). In response to these challenges, the Public Health Institute of Malawi (PHIM) established an active, real-time COVID-19 sentinel surveillance. This system was designed to complement routine surveillance by rapidly detecting changes in COVID-19 cases and allowing better characterization of COVID-19 and associated risk factors (7). The surveillance system objectives were to detect an increase in SARS-CoV-2 infections early, to characterize persons and locales at greater risk, and to identify possible reasons for increased risk. In this paper, we describe the trends in SARS-CoV-2 symptomatic and asymptomatic cases and their demographic and behavioral characteristics during the first six months of the COVID-19 sentinel surveillance in Malawi.

## Methods

### Sentinel surveillance design

This was a surveillance initiative that aimed at monitoring epidemiological and clinical trends in SARS-CoV-2 infection among sampled persons at sentinel sites in Malawi. The COVID-19 sentinel surveillance methodology was adapted from the WHO recommendations on integrated sentinel surveillance of influenza and SARS-CoV-2. (7)

### Sentinel surveillance locations

The sentinel surveillance was set up in July 2022 in seven purposively selected sites in seven districts: five HFs— Limbe Health Centre (Blantyre), Bwaila Hospital (Lilongwe), Matawale Health Centre (Zomba), Mzuzu Urban Health Centre (Mzimba North), and Mangochi District Hospital (Mangochi)— and two border PoEs—Mwanza border (Mwanza) and Songwe border (Karonga) (**Figure 1**). The HFs were selected from the districts with the highest number of reported SARS-CoV-2 infections and from facilities with a high volume of patients (8). The PoEs were selected due to their high-volume of incoming travelers from Mozambique and Tanzania, respectively.

**Figure 1:**
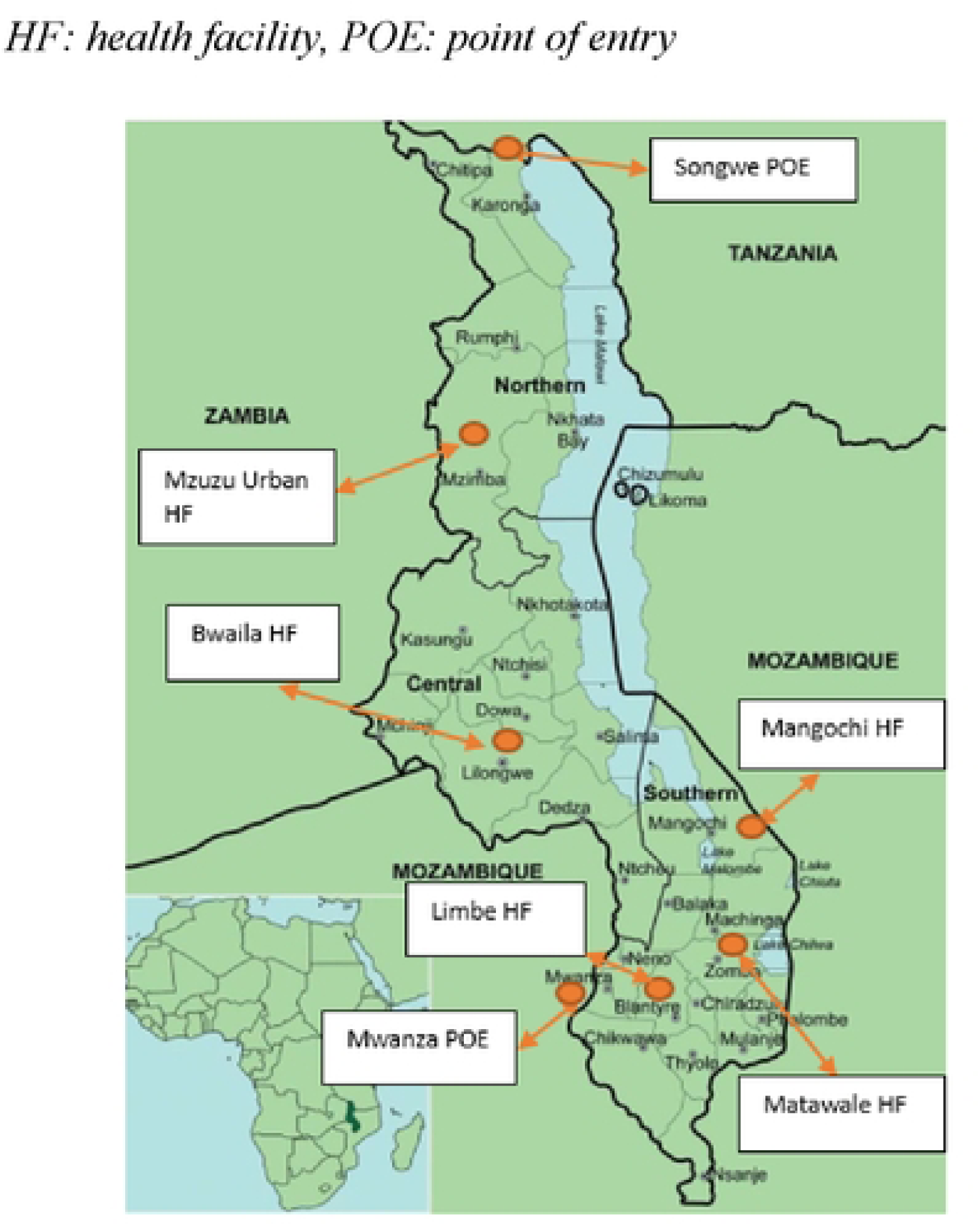
Map showing the seven COVID-19 sentinel sites across Malawi, July – December 2022.

### Study Populations

In HFs, the study population consisted of patients of any age seeking healthcare for any reason. We excluded severely ill patients, those under 18 years old without a legal guardian, or those who had previously participated in the survey within 14 days. Eligible participants were screened through clinical assessment and categorized into SARS-CoV-2 symptomatic or asymptomatic based on the WHO integrated COVID-19 and Influenza case definition. The symptomatic patients had symptoms of COVID-19/Influenza while asymptomatic group did not fulfil the case definitions^1^ (7). At the borders, persons ≥ 5 years of age entering the country were eligible to participate. In both populations, eligible participants or their guardians had to provide consent.

### Sample Size and Sampling strategy

The HF sample size calculation assumed a SARS-CoV-2 infection prevalence of approximately 2% within the general HF population based on unpublished results from previous local COVID-19 surveys. To ensure detection of at least one case of SARS-CoV-2 infection per week, given the 2% assumed prevalence and an estimated 10% refusal rate, each site aimed to recruit 50 symptomatic and 25 asymptomatic participants per week. This approach was consistent with WHO guidance on integrated SARS-CoV-2/Influenza surveillance (7).

Similarly, the sample size for the PoEs followed the same assumptions—a 2% infection prevalence and a 10% refusal rate— aiming to detect at least one SARS-CoV-2 case per week. Based on WHO technical guidance for SARS-CoV-2 surveillance, each PoE had an enrollment target of 50 travelers per week.

As part of sampling procedures, all participants in HFs were screened while entering the facility gate using an eligibility checklist and categorized as asymptomatic and symptomatic. From Mondays to Fridays, the first ten symptomatic and the first five asymptomatic eligible patients were then selected to participate through consecutive sampling.

For PoE, each week, a sampling interval was determined based on the estimated number of travelers crossing the border until a maximum of 50 participants were enrolled each week. A total of 10 travelers daily were selected from points of entry using systematic sampling, regardless of the presence of COVID-19 symptoms.

### Data and specimen collection

Before rolling out the implementation of the COVID-19 sentinel surveillance, all staff were trained on the protocol procedures. A dedicated surveillance assistant oversaw activities at each site and was responsible for data entry and quality control. Questions were administered using a structured questionnaire installed on an electronic tablet using the Open Data Kit (ODK) platform. The questionnaire was available in English, Chichewa, Tumbuka and KiSwahili. We collected self-reported information on socio-demographics, past exposure to COVID-19, current signs and symptoms of influenza-like illness, contact with PCR-confirmed SARS-CoV-2– infected persons, national and international travel, attendance to gatherings, adherence to COVID-19 preventive measures, underlying health conditions— HIV, cardiovascular disease, chronic respiratory disease, diabetes, and other medical conditions—, and vaccination against COVID-19.

A nasopharyngeal (NP) specimen was collected from all participants. The NP specimens were immediately put in viral transport media and transported in temperature regulated cooler boxes for storage at 2-8 °C before testing. NP specimen tests were done at the sentinel facilities utilizing RT-PCR GeneXpert platforms within 24 hours of collection. Specimens that tested positive for SARS-CoV-2 were stored in minus 80 degrees freezers within 72 hours of specimen collection prior to shipment to the PHIM national reference laboratory for genomic sequencing. All test results were incorporated into the routine surveillance system. The district health offices where the cases were detected were responsible for communicating the test results to the participants.

### Data management and analysis

Data collected was daily sent through internet connection to a secure server. The data were downloaded from the server and imported into Stata software, version 14.2 for data cleaning and analysis (9). Each participant was assigned a unique identifier upon enrollment for linking ODK data and laboratory results. If values exceeded clinically expected ranges, queries were sent to the concerned site to confirm their validity. Data from all the sentinel sites were assessed for consistency on a weekly basis. Outputs tables and graphs including weekly and cumulative enrolment figures, collected specimens, PCR results, and vaccination status for all participants were generated on a weekly basis and shared with the PHIM for investigation and action. COVID-19 vaccination status was self-reported by participants. Participants who reported two doses of COVID-19 AstraZeneca or Pfizer vaccine or one dose of COVID-19 Johnson & Johnson vaccine were categorized as “fully vaccinated” while those receiving one dose of COVID-19 AstraZeneca or Pfizer vaccine were categorized as “partially vaccinated”. Those without any dose of any vaccine were categorized as “not vaccinated at all”. We conducted descriptive [frequencies, medians, and interquartile ranges (IQRs)] and bivariate analyses (not included in this manuscript) of sociodemographic and clinical factors and associations with RT-PCR positivity rates (PR). We also compared PRs between symptomatic participants, asymptomatic participants, and travelers. Multivariable analyses assessed associations between PRs and various sociodemographic and clinical factors, adjusting for potential confounders: sex, age, education, time since last positive SARS-CoV-2 test, and contact with a positive SARS-CoV-2 case. Independent variables were occupation, travelling outside the country, COVID-19 vaccination status, time since last vaccination, underlying factors and pregnancy. We applied multivariable logistic regression (binary outcome was a positive or negative PCR result) incorporating covariates selected based on epidemiological and statistical importance from bivariate analyses (chi-square). Crude and adjusted odds ratios (aOR) and 95% confidence intervals (CIs) were calculated. We considered a P value ≤0.05 to be statistically significant.

### Ethical considerations

Ethics approvals were granted by the National Health Services Research Council (Reference NHSRC # 22/04/2896) and the protocol was reviewed by CDC, deemed not research, and was conducted consistent with applicable federal law and CDC policy (10,11).

All participants gave their written informed consent or assent prior to their participation in the surveillance procedures. Informed consent from a guardian was required for all children under 18 years of age, except for emancipated minors.

## Results

### Participant Screening and Enrollment

A total of 9,981 individuals were screened across all seven sentinel sites. Among them, 4.7% (n=471) were excluded from the study, either because they refused to give consent/assent (n=102) or were ineligible based on surveillance exclusion criteria (n=369). A total of 9,510 participants (95.3%) were enrolled;205 declined to provide NP specimens and were excluded. A total of 9,305 participants (93.2%) provided both the NP specimens and responded to the questionnaire (**Figure 2**).

**Figure 2:**
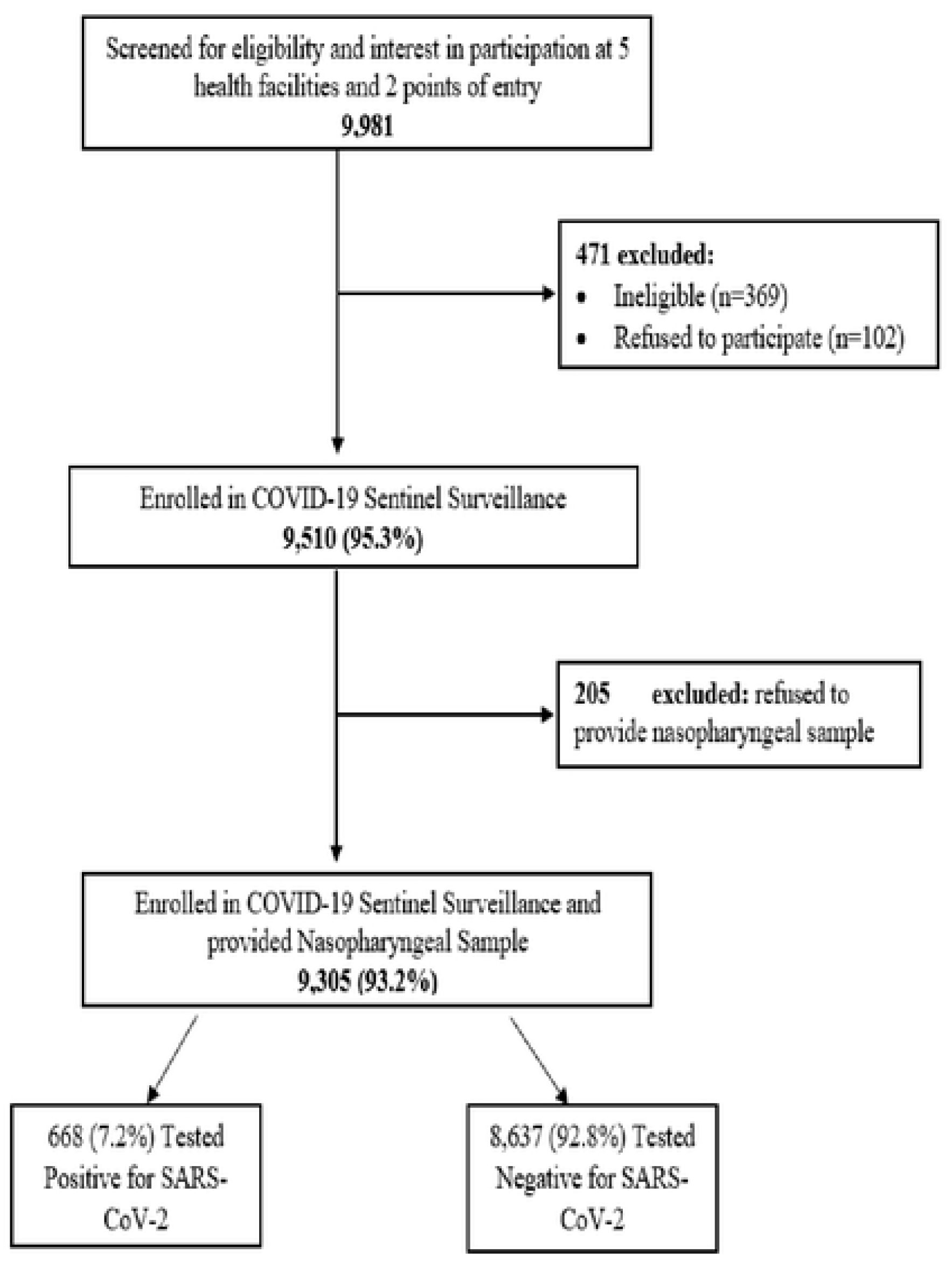
Flow diagram of participants inclusion, COVID-19 sentinel surveillance, Malawi, July - December 2022.

### Participant Characteristics by Sentinel Surveillance Site Type and Symptom Status

**Error! Reference source not found.** provides a summary of participant characteristics categorized by type of sentinel surveillance site and symptom status. In HFs, fewer participants were enrolled at Mzuzu and Limbe health centers (15.6% and 15.0% respectively). More participants were enrolled at Mwanza PoE (59.5%) compared to Songwe PoE (40.5%). Most of the respondents enrolled at the health facilities were aged between 15 to 29 years (45.9% among symptomatic and 43.9% among asymptomatic) and most enrolled travelers were aged 30-49 years (59.0%). At HFs, females represented 61.1% and 61.9% of asymptomatic and symptomatic groups, respectively. Among travelers, 70.7% were males. Most of the respondents had attended secondary education in both the HFs (72.7%) and PoEs (76.0%).

In the HFs, most of the respondents in both symptomatic and asymptomatic groups were “unemployed” (39.3% and 46.7% respectively). At PoEs, most participants were either employed or in business (46.5%).

About three quarters of the respondents in each category were not vaccinated:74.4% of symptomatic patients, 77.0% of asymptomatic patients and 70.3% of travelers. Less than one fifth in each category were fully vaccinated: 19.4% symptomatic, 19.9% asymptomatic and 18.1% of travelers.

Most respondents reported no underlying health condition (83.6%). Among the self-reported underlying conditions, HIV was the most reported: 5.3% of symptomatic patients, 10.8% of asymptomatic and 4.7% of travelers.

### COVID-19 Positivity Rate Trends among Health Facility Respondents and Travelers

**Figure 3** shows trends of COVID-19 positivity rate in those recruited at the HFs and the travelers. The highest overall positivity rate of 29.6% was recorded in the week of 11-15 July 2022 when the lowest number of participants were recruited (n=136) while the lowest positivity of 0.9% was recorded in the week of 24-28 October 2022 (n=334). The average overall positivity rate was 8.0%. The highest number of participants recruited was 531 in the week of 12-16 December 2022. On average 372 participants were recruited every week. Throughout the sentinel surveillance period, the positivity trends among the symptomatic and asymptomatic patients showed a similar pattern, with the symptomatic group showing slightly higher rates in most of the time points.

**Figure 3:**
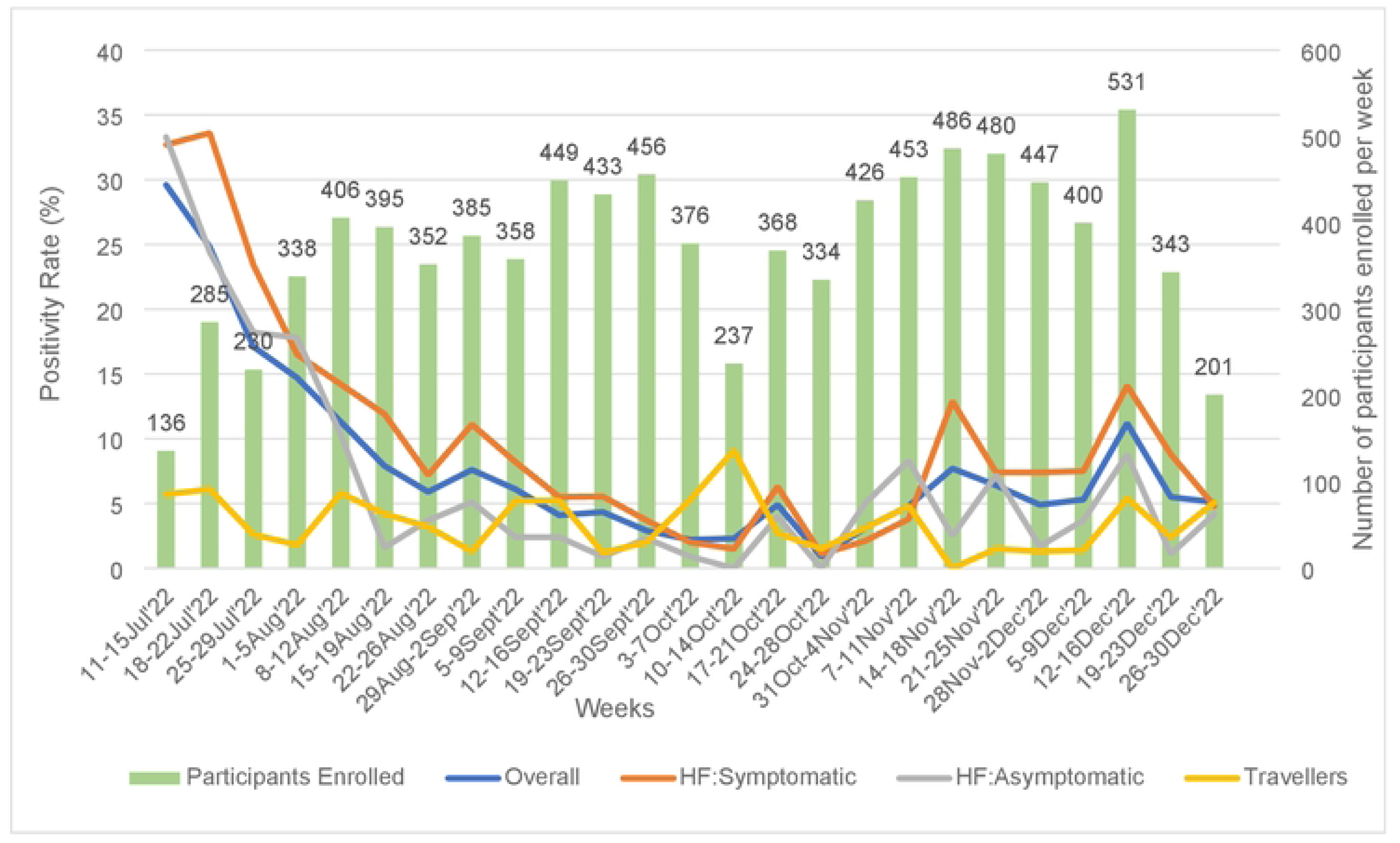
Trends of cumulative COVID-19 positivity rate, overall, among symptomatic and asymptomatic, and travelers, COVID-19 sentinel surveillance, Malawi, July-December 2022.

### Association of COVID-19 Positivity with Demographics and Behavioral Factors

**Table 2** presents the association of SARS-CoV-2 positivity rate with various demographic and behavioral factors categorized by symptomatic patients, asymptomatic patients, and travelers. The results of unadjusted results are only presented in the table. Below is a description of adjusted model results.

**Table 1:** Characteristics of enrolled participants by symptom status and type of sentinel surveillance site), COVID-19 sentinel surveillance, Malawi, July-December 2022.

|  |  | **Patients at health facilities** |  | **Travelers at points of entry (POEs)** | **Total study population** |
| --- | --- | --- | --- | --- | --- |
|  |  | **Symptomatic**<br>**N=4880** | **Asymptomatic**<br>**N= 2690** | **N=1735** | **N=9305** |
|  |  | **n (%)** | **n (%)** | **n (%)** | **n (%)** |
| **Characteristic** |  |  |  |  |  |
| **Sentinel Site** |  |  |  |  |  |
| Health Facilities | Mzuzu Urban | 1033 (21.2) | 150 (5.6) | NA | 1183 (15.6) |
|  | Bwaila | 1096 (22.5) | 655 (24.3) | NA | 1751 (23.1) |
|  | Mangochi | 1099 (22.5) | 740 (27.5) | NA | 1839 (24.3) |
|  | Matawale | 993 (20.3) | 670 (24.9) | NA | 1663 (22.0) |
|  | Limbe | 659 (13.5) | 475 (17.7) | NA | 1134(15.0) |
| POEs | Songwe | NA | NA | 703 (40.5) | 703 (40.5) |
|  | Mwanza | NA | NA | 1032 (59.5) | 1032 (59.5) |
| **Age (years)** | 0-14 | 236 (4.8) | 102 (3.8) | 41 (2.4) | 379 (4.1) |
|  | 15-29 | 2240 (45.9) | 1183 (43.9) | 514 (29.6) | 3937 (42.3) |
|  | 30-49 | 1858 (38.1) | 1104 (41.0) | 1024 (59.0) | 3986 (42.8) |
|  | ≥50 | 546 (11.2) | 301 (11.2) | 156 (9.0) | 1003 (10.8) |
| Median, years (IQR) |  | 29 (17) | 30 (16) | 34 (13) | 31 (17) |
| **Sex** | Female | 2981 (61.1) | 1664 (61.9) | 509 (29.3) | 5154 (55.4) |
|  | Male | 1899 (38.9) | 1026 (38.1) | 1226 (70.7) | 4151 (44.6) |
| **Highest level of education** | None | 316 (6.5) | 117 (4.4) | 11(0.6) | 444 (4.8) |
|  | Primary | 1089 (22.3) | 541 (20.1) | 405 (23.3) | 2035 (21.9) |
|  | Secondary | 3475 (71.2) | 2032 (75.5) | 1319 (76.0) | 6826 (73.3) |
| **Occupation** | In school | 706 (14.5) | 282 (10.5) | 137 (7.9) | 1125 (12.1) |
|  | Employed/Business | 1568 (32.1) | 793 (29.5) | 806 (46.5) | 3167 (34.0) |
|  | Farmer | 690 (14.1) | 358 (13.3) | 89 (5.1) | 1137 (12.2) |
|  | Unemployed | 1916 (39.3) | 1257 (46.7) | 703 (40.5) | 3876 (41.7) |
| **Vaccination Status** | Non-Vaccinated | 3631 (74.4) | 2070 (77.0) | 1220 (70.3) | 6921 (74.4) |
|  | Partially vaccinated | 303 (6.2) | 192 (7.1) | 199 (11.5) | 694 (7.5) |
|  | Full vaccination | 946 (19.4) | 428 (19.9) | 316 (18.2) | 1690 (18.1) |
| **Underlying Conditions** | HIV | 259 (5.3) | 290 (10.8) | 81 (4.7) | 630 (6.8) |
|  | CV diseases* | 180 (3.7) | 66 (2.5) | 42 (2.4) | 288 (3.1) |
|  | Chronic respiratory disease<sup>†</sup> | 204 (4.2) | 35 (1.3) | 21 (1.2) | 260 (2.8) |
|  | Diabetes | 30 (0.6) | 18 (0.7) | 10 (0.6) | 58 (0.6) |
|  | Others | 127 (2.6) | 80 (3.0) | 33 (1.9) | 240 (2.6) |
|  | None | 4080 (83.6) | 2201 (81.8) | 1548 (89.2) | 7829 (84.1) |

\*CV diseases: chronic cardiac disease and hypertension.
<sup>†</sup>Chronic respiratory disease: including asthma and tuberculosis.

**Table 2:**
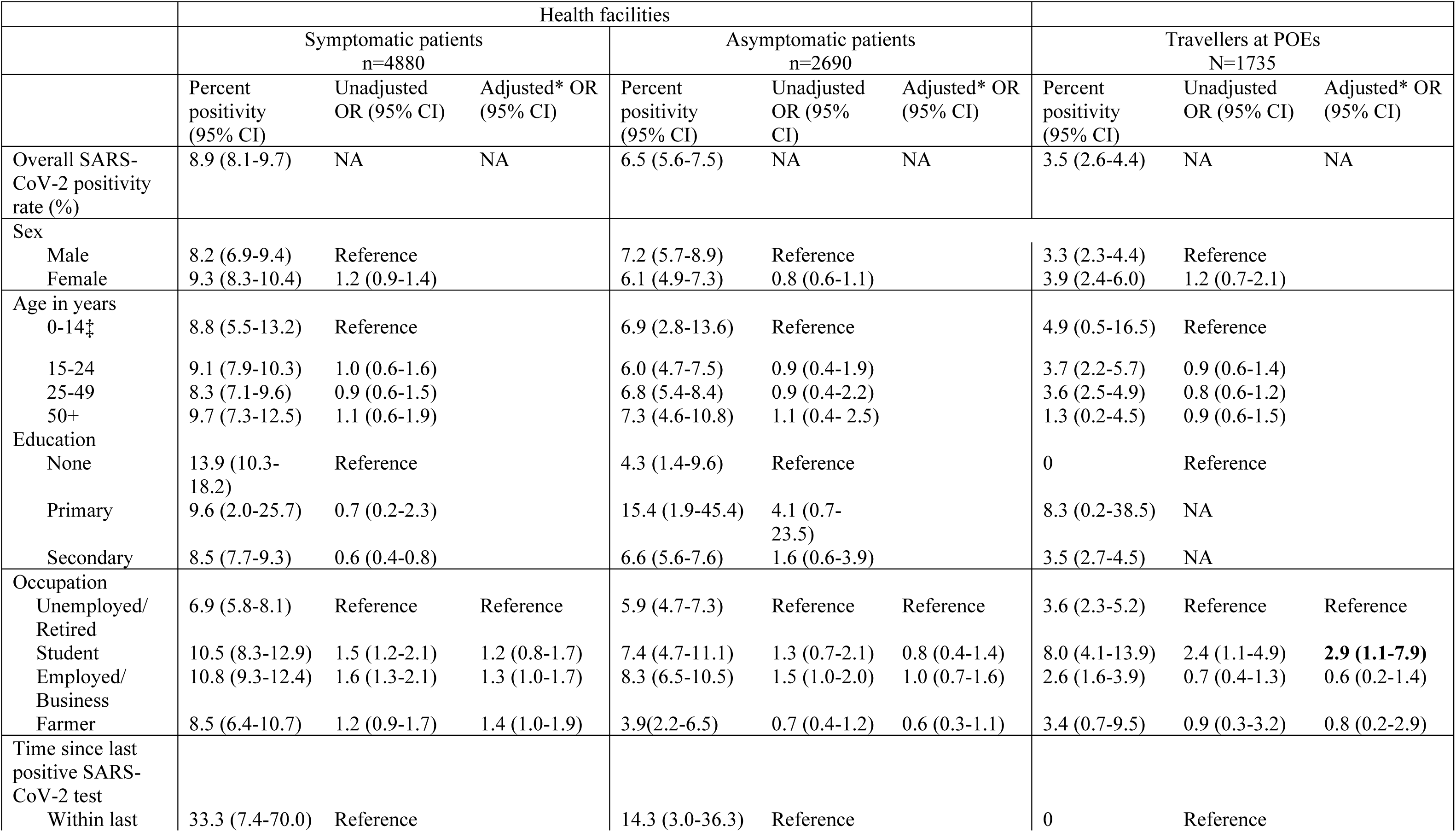
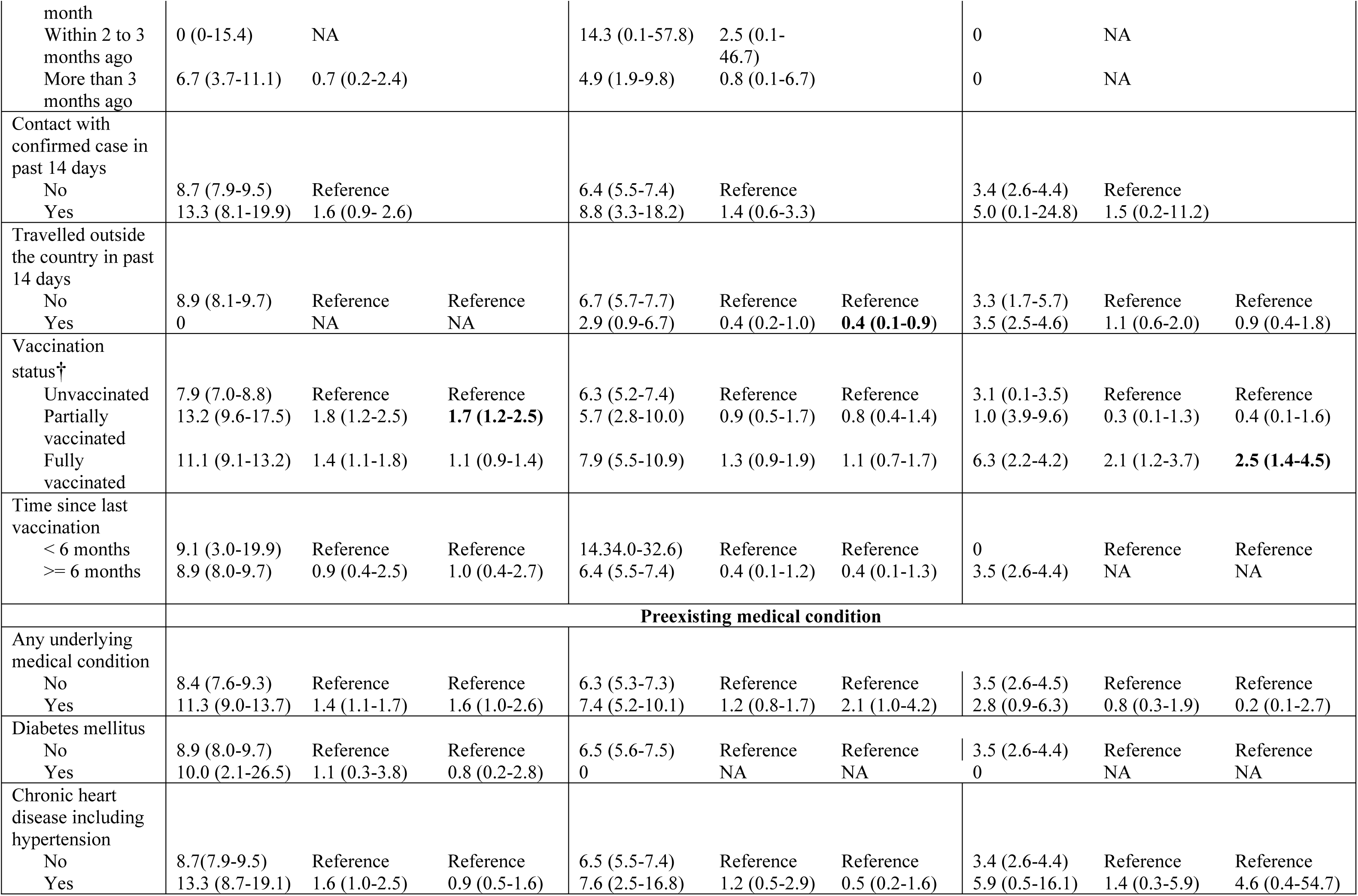
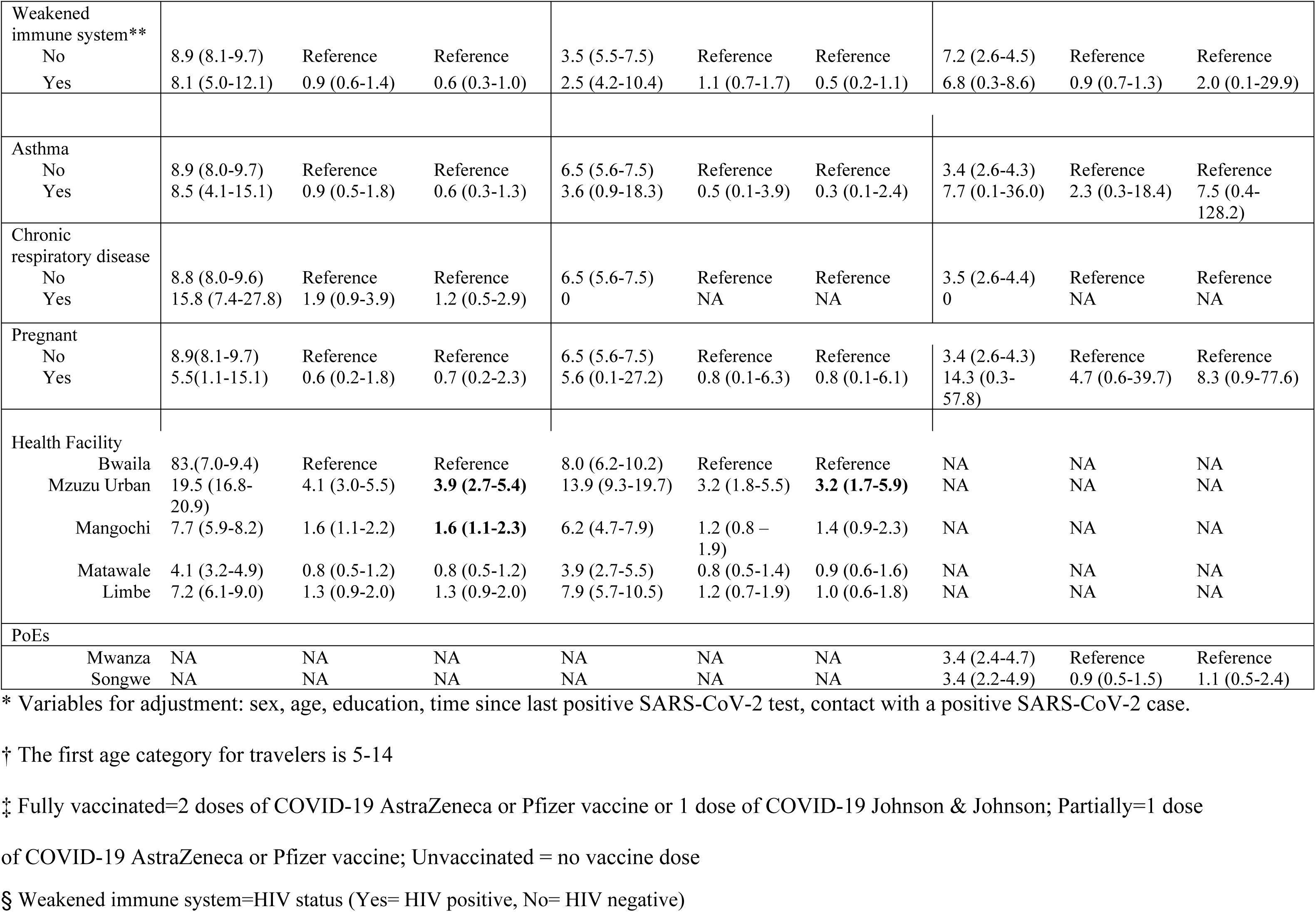
Association of SARS-CoV-2 positivity rates with demographic and behavioral factors, COVID-19 sentinel surveillance, Malawi, July-December 2022.

### Symptomatic patients

After adjusting for all potential confounders, those who were partially vaccinated remained more likely to test positive compared to those who were not vaccinated (aOR: 1.7; 95% CI: 1.2-2.5). Participants that enrolled at Mzuzu Urban and Mangochi health facilities were more likely to test positive to SARS-CoV-2 than those at the referenced facility (Mzuzu Urban: aOR:3.9; 95% CI:2.7-5.4; Mangochi: aOR: 1.6 95% CI: 1.1-2.3).

### Asymptomatic patients

In adjusted model, those who had travelled outside Malawi within the previous 14 days were less likely to test positive for SARS-CoV-2 compared to those that had not travelled (aOR: 0.4; 95% CI: 0.1-0.9). Those who enrolled at Mzuzu Urban health facility were more likely to test positive to SARS-CoV-2 than those at the referenced facility (aOR: 3.2; 95% CI: 1.7-5.9).

### Travelers

In adjusted model, students remained more likely to test positive for SARS-CoV-2 compared to those unemployed or retired (aOR: 2.9; 95% CI: 1.1-7.9). Travelers who were fully vaccinated were more likely to test positive for SARS-CoV-2 compared to those that were not vaccinated (aOR: 2.5; 95% CI: 1.4-4.5).

## Discussion

This sentinel surveillance was among the initial steps towards implementing integrated COVID-19 and Influenza surveillance in a resource limited setting (7). The sentinel surveillance offered an opportunity to re-train health workers on the need for continued active tracking of COVID-19 cases and routinize surveillance amidst other priority programs and emerging outbreaks. This in turn provided an opportunity for systems strengthening at all levels. The methodology used offered a unique possibility to evaluate the two different surveillance approaches within the same health system as the country prepared for a full integrated surveillance approach (7). Although the influenza component was not fully incorporated during this period, the ability to detect COVID-19 cases in both symptomatic and asymptomatic populations using the integrated COVID-19/Influenza case definition means that, to some extent, integration would be a feasible and potentially efficient approach, particularly in resource limited countries. On the contrary, the routine system alone could easily miss asymptomatic patients particularly in the Malawi settings where the COVID-19 testing targeted those with symptoms. (12)

This sentinel surveillance helped bridge some gaps of inadequate SARS-CoV-2 testing, as some routine testing sites in Malawi were not conducting tests at all according to unpublished data from daily COVID-19 updates. The low testing at some routine sites might have been due to the general relaxation of COVID-19 control measures, which is commonly observed during low incidence periods when preventative communications and other measures tend to be minimal (13). The other reason for low testing could be the shifting of attention of surveillance efforts to other emergencies at the time, specifically the national-wide cholera outbreak that begun in March 2022. (5)

The difference in sex distribution among participants enrolled from the HFs as compared to the PoEs might reflect some known differences in gender and childbearing roles. Due to their roles in childbearing and primary caregiving for children, women go to health facilities more often than men. On the other hand, men, due to their perceived primary roles as home providers, are often more engaged than females in travelling for businesses (14).

The trend showed a relatively higher positivity rate at the beginning of the surveillance period, which may be an overestimation due to the smaller sample size during that early phase.” (15). The rest of the epidemiological curve from the COVID-19 sentinel surveillance was comparable to the overall curve inclusive of all other cases tested in the routine surveillance sites (16). This epidemiology curve, which was relatively flat compared to the previous year, aligns with the predictions made by Jiang F and colleagues based on their statistical simulation models. They suggested that this flattening is probably a result of the combined effect of various preventative measures, including immunizations, as control strategies in most countries became more systematic. (17)

In this surveillance, the similarity in the trends of positivity rate among symptomatic and asymptomatic population highlight the need for targeting both populations for testing and preventative measures Asymptomatic SARS-CoV-2 cases can be targeted through various strategies, such as testing those who had contact with confirmed cases and incorporating asymptomatic testing into integrated surveillance by deliberately sampling asymptomatic patients on a weekly basis (18).

Various factors were evaluated to be associated with SARS-CoV-2 positivity rate in this surveillance. The positive association for students and employees may be explained by their increased risk, as these occupations are more likely to involve physical contact than those who are not employed. Similarly, Mangochi hospital and Mzuzu urban health centre showed a higher likelihood of a SARS-CoV-2 positivity rate as compared to Bwaila urban in Lilongwe city. This could be due to a complex interplay of different socio-economic factors such as level of education and access to health preventative services considering the differences in levels of urbanization in these districts (*17,19*). These in-country district level differences are relevant for prioritizing resources in resource limited settings. Asymptomatic HFs participants who had traveled outside the country within the past 14 days were less likely to test positive to SARS-CoV-2. Travelling history might have lowered their overall risk through mandatory screening procedures and awareness campaigns that were in place in many countries’ PoEs (20).

This surveillance found that those who were partially vaccinated among the symptomatic group in HFs and the travelers who were fully vaccinated were more likely to test positive to SARS-CoV-2 compared to the unvaccinated. This association was not observed in the asymptomatic group in HFs for both partially and fully vaccinated categories, and among travelers who were partially vaccinated. These inconsistent findings across the categories need to be interpreted with caution considering some limitations of this surveillance. To start with, COVID-19 vaccination status was self-reported, thus subject to reporting and misclassification biases. Secondly, the smaller numbers of those that were fully/partially vaccinated, can result in less precise estimates of effect sizes. Therefore, given these inconsistent findings across the categories, the complexity of the issue and the limitations of this study, we would need further evaluations to explain the possible reasons for this association. Suffice to say that the evidence of COVID-19 vaccination efficacy and effectiveness is well-documented, particularly for preventing symptomatic disease (21,22). As a recommendation, there is need for intentionally designed surveillances to incorporate some behavioral issues that may impact COVID-19 vaccination effectiveness at population level. Additionally, an important phenomenon that should be kept in check is the so-called “Peltzman Effect”. This refers to a behavioral compensation mechanism whereby individuals would not adhere to precautionary measures due to the perceived protection from vaccine (23). This phenomenon, if at work within a population, would result in increased COVID-19 transmission in the vaccinated population that does not comply to preventative measures.

In general, this surveillance had some limitations. The results do not represent the entire population as this was limited to a defined high-risk population that would differ in some epidemiological factors. Additionally, considering the nature of the desired population, the sampling methodology in the health facilities could not be left to systematic sampling, as was in our first protocol design, as that would affect the sample size as was shown during piloting. The use of consecutive sampling for both symptomatic and asymptomatic groups means that selection bias cannot be excluded. Lastly, our questionnaire did not collect data on some other key areas, such as disease severity and COVID-19 vaccination behavioral questions.

## Conclusion

This COVID-19 active sentinel surveillance complemented routine passive surveillance and was instrumental in the low incidence period. The surveillance offered lessons to feed into a full integration approach of COVID-19 and Influenza surveillance in Malawi as per WHO recommendations. The lower observed prevalence of SARS-CoV-2 and the relatively flat trend imply that some preventative measures and systems were in place and working. The ongoing detection of cases in both symptomatic and asymptomatic groups, however, underscores the important need of continuous COVID-19 surveillance in all populations. Moreover, the ongoing detection of infections among travelers, despite instituted COVID-19 travel restrictions, indicates the need for continued surveillance and provides an opportunity to detect novel variants coming into the country. The results also underscore the need for in-depth study of the interaction between COVID-19 vaccinations and behavioral adherence to preventative measures so that sensitization messages can be formulated according to context and evidence.

## Data Availability

All data underlying the findings reported in the manuscript are deposited in repository managed by Ministry of Health-Malawi a public repository

## Acknowledgments

The authors thank the people of Malawi and the international organizations that contributed to this work.

## Disclaimer

The findings and conclusions in this manuscript are those of the authors and do not necessarily represent the official position of the funding agency.

## Author Biography

Godwin Ulaya (MPH, MBBS) led the COVID-19 Surveillance initiative at I-TECH Malawi and Public Health Institute of Malawi (PHIM). As a Project manager, his role involved mobilizing various Stakeholders in responding to the pandemic. With an interest in epidemiological research and management, he has been involved in various health programs under Ministry of Health, Malawi and in Clinical trials whilst at Johns Hopkins Research Project.

## Notes

### Competing Interest Statement

The authors have declared no competing interest.

### Funding Statement

This survey was supported by the President's Emergency Plan for AIDS Relief (PEPFAR) with CARES Act funding through the US Centers for Disease Control and Prevention (CDC) under the terms of Cooperative Agreement NU2GGH002038 with implementation led by International Training and Education Center for Health (I-TECH) Malawi, University of Washington, Lilongwe, Malawi The findings and conclusions in this manuscript are those of the authors and do not necessarily represent the official position of the funding agency.

### Author Declarations

Ethics approvals were granted by the National Health Services Research Council (Reference NHSRC # 22/04/2896) and the protocol was reviewed by CDC, deemed not research, and was conducted consistent with applicable federal law and CDC policy

